# Coincident rapid expansion of two SARS-CoV-2 lineages with enhanced infectivity in Nigeria

**DOI:** 10.1101/2021.04.09.21255206

**Authors:** Egon A. Ozer, Lacy M. Simons, Olubusuyi M. Adewumi, Adeola A. Fowotade, Ewean C. Omoruyi, Johnson A. Adeniji, Taylor J. Dean, Janet Zayas, Pavan P. Bhimalli, Michelle K. Ash, Adam Godzik, Jeffrey R. Schneider, João I. Mamede, Babafemi O. Taiwo, Judd F. Hultquist, Ramon Lorenzo-Redondo

## Abstract

The emergence of new SARS-CoV-2 variants with enhanced transmissibility or decreased susceptibility to immune responses is a major threat to global efforts to end the coronavirus disease 2019 (COVID-19) pandemic. Disparities in viral genomic surveillance capabilities and efforts have resulted in gaps in our understanding of the viral population dynamics across the globe. Nigeria, despite having the largest population of any nation in Africa, has had relatively little SARS-CoV-2 sequence data made publicly available. Here we report the whole-genome sequences of 74 SARS-CoV-2 isolates collected from individuals in Oyo State, Nigeria in January 2021. Most isolates belonged to either the B.1.1.7 Alpha “variant of concern” or the B.1.525 Eta lineage, which is currently considered a “variant of interest” containing multiple spike protein mutations previously associated with enhanced transmissibility and possible immune escape. Nigeria has the highest reported frequency of the B.1.525 lineage globally with phylogenetic characteristics consistent with a recent monophyletic origin and rapid expansion. Spike protein from the B.1.525 lineage displayed both increased infectivity and decreased neutralization by convalescent sera compared to Spike proteins from other clades. These results, along with indications that the virus is outpacing the B.1.1.7 lineage in Nigeria, suggest that the B.1.525 lineage represents another “variant of concern” and further underline the importance of genomic surveillance in undersampled regions across the globe.

## INTRODUCTION

A little over a year after its emergence in the Hubei province of China, the continued spread of severe acute respiratory syndrome coronavirus 2 (SARS-CoV-2) across the globe has sparked a worldwide health crisis ^1-3^. Despite the relatively low mutation rate of this virus, its high prevalence in the human population globally has allowed it to diversify quickly ^4,5^. Identification and tracking of these mutations through whole genome sequencing efforts have been critical to identifying routes of transmission, mapping outbreaks across communities over time, and characterizing new variants that may change the virological or clinical aspects of the disease ^6-9^. For example, during the spring and summer of 2020, a mutation in the viral Spike protein, D614G, was identified in association with higher viral loads in patient upper airways ^10,11^. The rapid expansion of this variant in communities across the world coupled with virological assays *in vitro* and *in vivo*, suggest that this variant is more transmissible than earlier lineages ^12-15^. Indeed, this mutation is nearly fixed in the global SARS-CoV-2 population today ^11^.

Continued surveillance efforts have since identified a number of ‘variants of concern’ that have been associated with increased transmissibility, increased disease severity, decreased susceptibility to therapeutic agents, or decreased susceptibility to antibody neutralization^16^. Using the dynamic Pangolin nomenclature for SARS-CoV-2 (cov-lineages.org), these include the B.1.1.7 lineage originally identified in the United Kingdom, the B.1.351 variant originally identified in South Africa, and the P.1 variant originally identified in Brazil ^17-22^. One of the major concerns is that these viruses may prove to be more transmissible and could result in current public health standards being less effective at containing spread of the virus ^23-25^. Furthermore, these changes to the viral Spike protein may render current vaccine formulations less efficacious and/or confer increased capacity for re-infection ^26-28^. Additionally, viral mutations may impact the effectiveness of therapeutic monoclonocal antibodies or other antiviral treatments ^29^. Finally, there is the possibility that some changes may be associated with increased disease severity. Viral lineages with specific mutations that are predicted to affect transmission, diagnostics, therapeutics or immune escape but for which these effects have not been demonstrated are classified as “variants of interest” ^30^. Current variants of interest include the B.1.525, B1.526, B1.617 (among which B.1.617.2 has been upgraded to “variant of concern” ^16^), and P2 lineages. Addressing the threats posed by variants of concern and variants of interest requires continued surveillance of SARS-CoV-2 genetic diversity worldwide and assessment of the impact novel mutations or combinations of mutations in emerging lineages may have on viral fitness, transmission, antibody neutralization, therapeutics, and pathogenesis.

While many countries have developed extensive genetic surveillance and reporting systems, several regions across the globe remain critically undersampled ^31^. For example, several countries in Africa have reported only a handful of sequences relative to their cumulative case counts, if any ^32,33^. Significantly, Nigeria, which has the largest population of any African country, has only a small number of reported SARS-CoV-2 sequences.

Through mid-February of 2021, the majority of the approximately 300 SARS-CoV-2 genome sequences publicly available from Nigeria had been generated through the African Centre of Excellence for Genomics of Infectious Disease (ACEGID), which also serves other African countries (www.acegid.org). However, given Nigeria’s status as an epicenter of commerce and travel in Africa, undetected expansion of a more infectious, virulent or immune-escape variant in Nigeria could have major repercussions for the entire continent. More consistent and higher volume of sample collection and sequencing is required in Nigeria to strengthen the public health value of COVID-19 surveillance efforts.

To better understand the SARS-CoV-2 population structure in Nigeria and add samples from this region, we set out to perform whole genome sequencing of SARS-CoV-2 viruses originating in Oyo state, Nigeria. SARS-CoV-2 sequencing was performed on 74 nasopharyngeal specimens collected from COVID-19 patients in Oyo state in January 2021. Phylogenetic analysis revealed two primary lineages of virus circulating in the region: B.1.1.7, or Alpha, a global ‘variant of concern’, and B.1.525, or Eta, currently designated a ‘variant of interest’. We show that isolates from both lineages are largely monophyletic, suggesting singular introductions sometime in the fall of 2020. Using a pseudotyped virus system, we found that mutations in the Spike protein gene of B.1.525 lineage isolates promoted increased viral entry into cells expressing the entry receptor ACE2. Furthermore, they decreased the effectiveness of antibodies from naturally-infected individuals to neutralize the virus, though antibodies from individuals immunized with mRNA vaccines were still effective. Together, these data add to SARS-CoV-2 genomic surveillance in Nigeria, identify two lineages of note that dominated the epidemic in the region, and suggest that the B.1.525 lineage may represent a currently underappreciated variant of concern.

## RESULTS

### Specimen characteristics

We obtained 80 specimens from COVID-19 patients in Oyo state, Nigeria collected between January 2 and January 14, 2021. These specimens consisted of residual nasopharyngeal and oropharyngeal swabs that tested positive for SARS-CoV-2 by quantitative reverse transcriptase PCR (qRT-PCR) diagnostic testing in the Biorepository and Clinical Virology Laboratory at the University College Hospital, College of Medicine at the University of Ibadan. The qRT-PCR cycle threshold (Ct) values of these specimens obtained in the clinical laboratory ranged from 13.46 to 33.11 for the N target (either DaAn Gene or BGI detection kits) and 18.51 to 37.10 for the ORF1ab target (DaAn Gene kit). The average age of the patients was 42 years (range 11-84) and the sex distribution was 50% female and 50% male. RNA was extracted from each sample and the presence of viral RNA confirmed by one-step qRT-PCR (CDC assay, RNaseP and N1 primer set) ^34^. RNA samples of sufficient quality (RNaseP control, Ct value <35) and with sufficient copies of the viral genome for sequencing (N1, Ct value <32) were reverse transcribed into complementary DNA (cDNA). The SARS-CoV-2 genome was subsequently amplified by multiplex PCR using the ARTIC protocol (primer set version 3) and subjected to deep sequencing on the Illumina platform ^35,36^. The minimum threshold for base calling was 10 reads with 90% coverage required across the genome to report the whole genome sequence. Of the 80 specimens, 6 failed to yield a satisfactory consensus sequence due to insufficient genetic material, insufficient purity after barcoding, or inadequate read coverage after sequencing; these samples were excluded from further analysis. The final 74 complete SARS-CoV-2 genomes were deposited in the public GISAID database (**Supplemental Table 1**) and subjected to phylogenetic analysis ^37,38^.

### The two dominant lineages in Oyo state are the lineage of concern B.1.1.7 and the emerging lineage of interest B.1.525

We first performed phylogenetic reconstruction using maximum likelihood (ML) phylogenetic analysis of the 74 SARS-CoV-2 genomes in this study using IQ-Tree v2.0.5. The dates of sample collection were subsequently integrated to build a temporal tree using TreeTime v0.7.6 (**Figure 1**). Most of the specimens from Oyo state (88%) belonged to one of two main clusters with very strong support at their base nodes (support: >97% aLRT and >97% UFboot). We used the Pango classification scheme to identify the lineages of the major clusters. The most abundant lineage (46 out of 74 sequences) was B.1.1.7. This lineage of concern, now referred to as the Alpha lineage by the World Health Organization, is associated with the Spike N501Y and other mutations and was first identified in the U.K. in the fall of 2020 ^20^. The second most common lineage (17 of 74 sequences) was a lineage of interest, designated B.1.525 by Pango or Eta by the WHO. This lineage contains the Spike E484K, Q677H, and F888L mutations as well as three in-frame deletions shared with B.1.1.7, including Spike 69-70del and Spike 144del (cov-lineages.org). The remaining sequences all belonged to other common B.1 lineages, except for 2 sequences from lineage A. Viruses from lineage A viruses spread throughout the world early in the pandemic, but are now relatively rare globally, accounting for only 0.3% of isolates sequenced and deposited in January 2021 (nextstrain.org).

**Figure 1.**
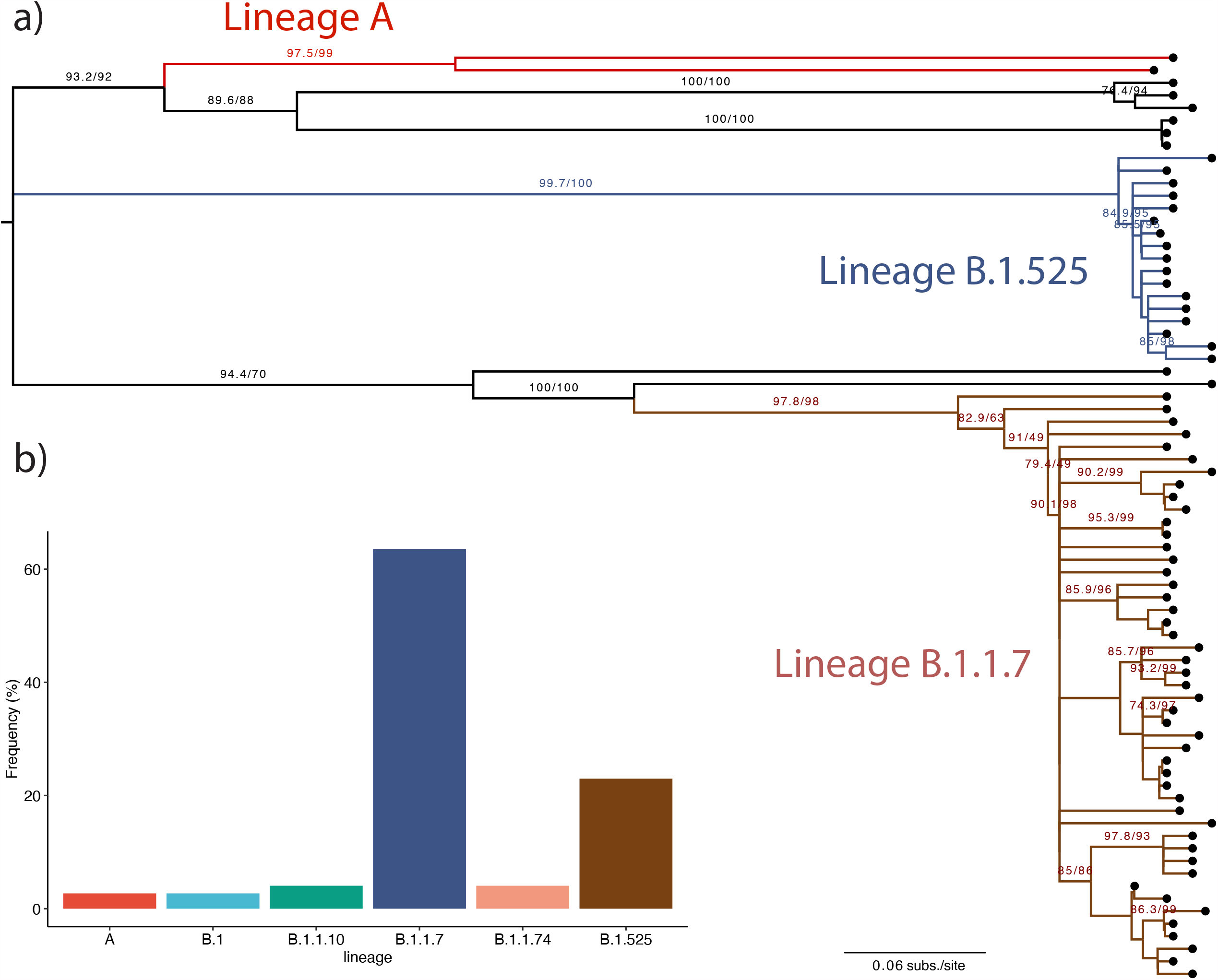
Phylogenetic analysis of SARS-CoV-2 isolates in Oyo state. **a)** ML phylogenetic tree of 74 SARS-CoV-2 specimen genomes in Oyo state collected between January 2 and January 14, 2021. All non-zero statistical support values for each branch are indicated. Lineages of interest are indicated and colored. Midpoint rooting was used for representation purposes. **b)** Distribution of the different pangolin lineages found in the Oyo dataset reported here.

To better determine if these sequences in Oyo state were representative of Nigeria as a whole, we downloaded all available sequences sourced from Nigeria in the GISAID database as of February 14^th^ (266 sequences, excluding those reported above). We followed the same approach as above and performed ML phylogenetic reconstruction to estimate the evolutionary relationships with the other available sequences. We also performed ancestral reconstruction of the most likely sequences at internal nodes as well as at transition points between geographical locations to better examine relationships between the isolates found in different states in Nigeria. This analysis confirmed a distribution of the viral populations throughout the country similar to what was observed in Oyo state. Of all reported sequences between January 1, 2021 and February 14, 2021, we observed that 68% of the sequences belonged to the B.1.1.7 lineage while 17% belonged to the B.1.525 lineage (**Supplementary Fig. 1**). This includes sequences from Lagos state, Osun state, Abuja, and other sequences from Oyo state collected outside this study. Overall, this result suggests that the distribution of lineages observed in this study is broadly reflective of the contemporaneous distribution of lineages in Nigeria and confirms the recent increase in prevalence of these two lineages coincident with an increase in overall case counts in the country ^39^.

### The B.1.1.7 and B.1.525 lineages in Nigeria are largely monophyletic

To better understand these data in the context of the global epidemic, we selected 4000 randomly sampled global sequences from GISAID collected between January 22^nd^, 2020 and January 26^th^, 2021 and repeated these analyses. We confirmed the clustering of the lineages as observed above (**Figure 2**). Furthermore, these analyses found that most B.1.1.7 lineage isolates in Nigeria were monophyletic, suggesting either a single successful introduction or multiple, closely linked introductions compatible with the observed founder effect. To confirm the B.1.1.7 sub-clustering in Nigeria, we performed a ML phylogenetic reconstruction of all B.1.1.7 sequences obtained in this study alongside 4000 randomly sampled B.1.1.7 sequences from across the globe (**Supplementary Fig. 2**). Additionally, we performed Bayesian inference of all B.1.1.7 sequences available from Nigeria with a smaller set of 500 randomly sampled B.1.1.7 sequences from across the globe (**Supplementary Fig. 3**). These analyses both confirmed strong support for a monophyletic sub-cluster that included almost all of the B.1.1.7 sequences from this study. Given these results, we estimated a time to the most recent common ancestor (TMRCA) for this B.1.1.7 sub-cluster to be October 28, 2020 [95% Highest Posterior Density (HPD) interval: October 16 - November 3, 2020].

**Figure 2.**
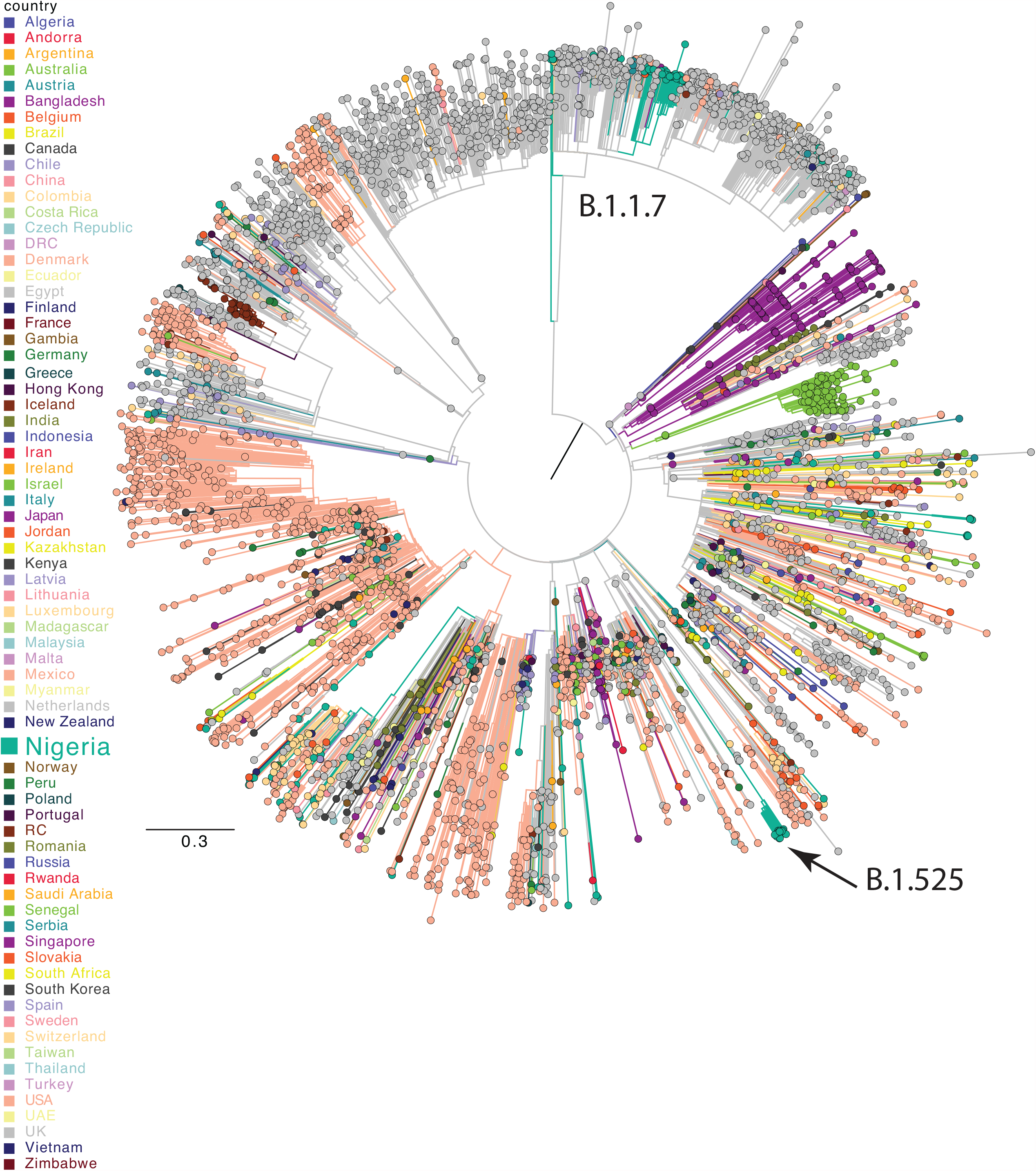
Phylogenetic analysis of Nigerian sequences compared to the global pandemic. ML phylogenetic temporal reconstruction of full genome sequences from Nigeria and 4000 randomly sampled global sequences from GISAID as of February 14^th^, 2021. Clades corresponding to B.1.1.7 and B.1.525 lineages are indicated. Branches and tips are colored by country.

The second most prevalent lineage observed in our dataset, B.1.525, is not yet highly prevalent on a global scale, but was recently defined as a lineage of international significance ^40^. As of February 14, 2021, only 159 sequences were available in GISAID from this lineage, of which Nigeria was the most frequent country of origin, accounting for roughly 25% of sequences ^40^. Referring to our previous ML analysis of the sequences from Oyo state alongside 4000 randomly selected global sequences (**Figure 2**), we observe strong support for a monophyletic clade of the B.1.525 lineage in Nigeria, suggestive of a single introduction followed by rapid expansion (**Figure 2**). Using a Bayesian approach, we next analyzed all 159 B.1.525 sequences available in GISAID alongside a smaller subset of B.1 sequences to help root the tree (the only 2 B.1 sequences present among the Oyo state isolates and 10 randomly selected from GISAID). The estimated TMRCA was October 27, 2020 [95% HPD interval: September 27 - November 21, 2020], similar to the TMRCA estimated for the B.1.1.7 lineage. Together, these analyses suggest that this lineage may have expanded principally in Nigeria, though additional sampling in West Africa and elsewhere is needed (**Figure 3**). Taken together, these analyses are all consistent with a single introduction or multiple, closely linked introductions of both the B.1.1.7 and B.1.525 lineages into Nigeria in the fall of 2020. These introductions were followed by expansion of each population to become the predominant two lineages in the country by early 2021.

**Figure 3.**
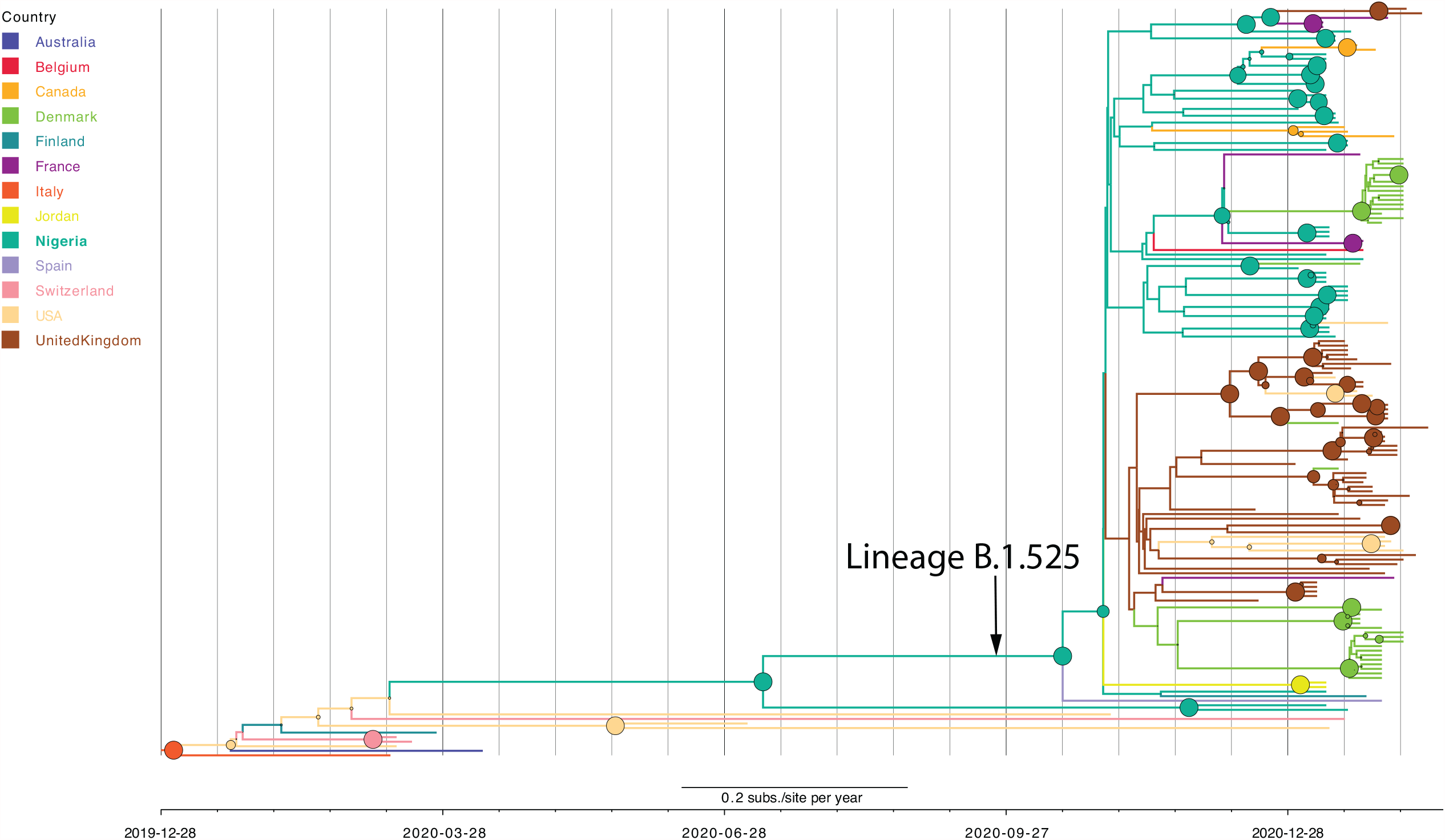
Phylodynamic tree of the entire B.1.525 lineage. Maximum clade credibility tree where branch colors represent the most probable geographical location of their descendent node inferred through Bayesian reconstruction of the ancestral state. All full genome B.1.525 sequences from this study and from GISAID as of February 14th, 2021 were included in the analysis. We included two B.1 lineage sequences from Nigeria and 10 randomly sampled from GISAID to help root the phylogenies. The branch that leads to the node that represents the B.1.525 MRCA is indicated. The width of the node circles represents their posterior probability.

### No significant differences are observed in the Ct values of the diagnostic PCR tests by lineage

Next, we compared the clinical Ct values measured at the time of diagnosis to assess for differences in viral load in patient upper airways by lineage. Using a linear model to control for patient age and sex, we compared the Ct value measured for the N1 probe for all 74 diagnostic specimens batched by lineage (**Supplementary Fig. 4**). We did not observe any statistically significant differences between the two main lineages (B.1.1.7 and B.1.525), and the remaining lineages lacked sufficient representation for meaningful comparison. Although a larger dataset that controls for a higher number of confounders will be needed to robustly test for differences, this result suggests there are comparable average viral loads in the upper respiratory tract between these two lineages at the time of patient diagnosis. B.1.1.7 has been previously associated with increased viral loads relative to other lineages ^41^, but those are not represented in sufficient numbers here for comparison.

### Spike protein mutations in the B.1.525 lineage promote cell entry

Given that the expansion of B.1.525 lineage isolates in Nigeria occurred alongside expansion of the known B.1.1.7 variant of concern, we sought to characterize the phenotypic impact of B.1.525 Spike (S) mutations on viral infectivity. B.1.525 lineage isolates are characterized by several nonsynonymous mutations in S at positions Q52R, A67V, E484K, D614G, Q677H, and F888L, as well as by two in-frame deletions at positions 69-70 and 144. Notably, both deletions are also found in the B.1.1.7 lineage and have been previously linked with enhanced infectivity and transmission ^42^, while the E484K mutation has arisen independently in multiple lineages and has been associated with potential for immune evasion ^43^.

Mapping the B.1.525 lineage-defining mutations onto available Spike protein structures demonstrated an accumulation of changes in the N-terminal domain (NTD) of the protein. To better understand the impact of these mutations, we used structural modeling to examine the predicted changes in Spike protein structure in the presence and absence of binding to both ACE2 as well as NTD-directed antibody (**Figure 4a**). As suggested by studies in other lineages, the E484K mutation in the receptor-binding domain (RBD) modifies a main epitope for class 2 antibodies. In addition, several other mutations in the B.1.525 lineage are immediately adjacent to another antigenic “supersite” located in the NTD ^44-46^, independent of the RBD. Deletion of the residue Y144 in particular directly impacts the central ‘N3 loop’ in the supersite while the 69-70 deletion and the A67V mutation further shift this loop. The Q52R mutation is also located in the NTD, but further from the identified antigenic region. Q677H is close to the furin cleavage site, potentially affecting the dynamics of Spike cleavage, a process required for cell entry ^47^.

**Figure 4.**
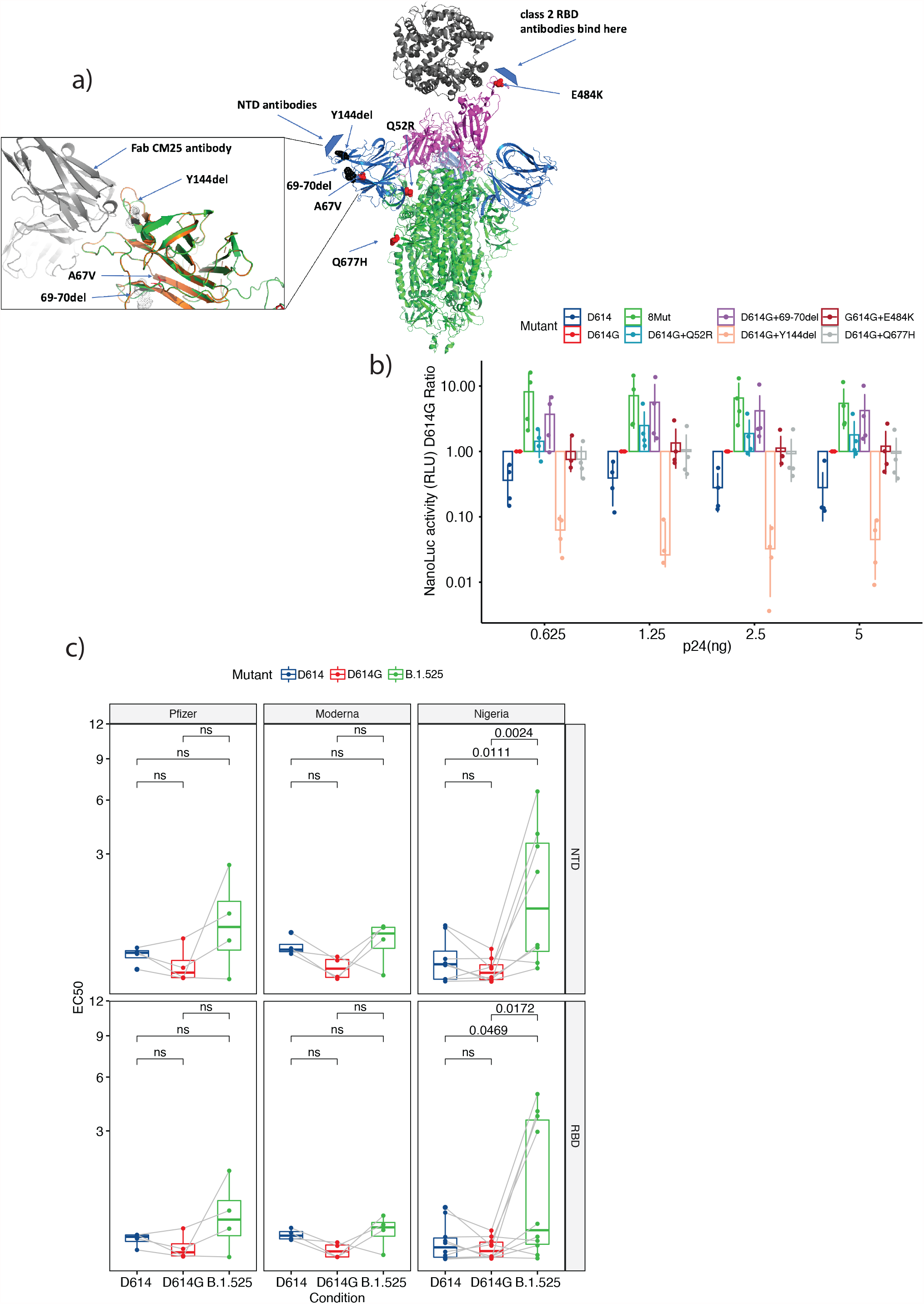
Analysis of B.1.525 Spike mutations on cellular entry and antibody neutralization. a) Overall structure of the SARS-CoV-2 spike protein trimer with N-terminal domains (NTD) in blue, receptor binding domains (RBD) in magenta, and ACE2 receptor in dark grey, based on the set of PDB coordinates 7a94 ^80^ showing the spike protein bound to one ACE2 molecule. Mutated residues in the B.1.525 spike are shown in chain A as red spheres, deleted residues as black spheres. A model of the B.1.525 NTD in the inset is shown interacting with the Fab C25 antibody (PDB 7m8j) ^77^. The unmodified chain on the wild type NTD is shown in orange. A significant conformational change in the N3 loop results in a significant drop in the (estimated) binding energy to this antibody, from -10.8 kcal/mol to -1.3. b) Nanoluc activity measured in relative light units (RLU) ratio between each of the mutants tested and the D614G mutant. D614G was used to calculate the ratios due to its predominance in the population before the appearance of the B.1.1.7 and B.1.525 lineages. Values are shown for each of the dilutions used after p24 concentration normalization. Bars represent the mean and lines represent the standard deviation of the replicates. c) Neutralization EC_50_ comparison between the different Spikes tested in the presence of sera from vaccinated (Pfizer or Moderna groups) or naturally-infected (Nigeria group) individuals. EC_50_ values were estimated using a four-parameter log-logistic function either with the NTD or RBD antibody concentrations. Statistically significant FDR values are indicated for within mutant comparisons (ns indicates non-significant FDR).

To test the impact of these mutations on viral entry, we generated a panel of HIV-1 luciferase reporter viruses pseudotyped with SARS-CoV-2 Spike containing selected mutations. Given that the Spike D614G mutation is nearly fixed on the global scale, this construct was used as a baseline. On this background, each of the other 8 mutations in B.1.525 were introduced both individually (with the exception of A67V that was initially not considered as a lineage-defining mutation and F888L that we were not able to grow) as well as cumulatively to generate a B.1.525 consensus Spike. The ancestral SARS-CoV-2 Spike D614 was also included as a control. Viral input was normalized by concentration of the HIV p24 protein and used to challenge HeLa cells overexpressing ACE2 in step-dilution MOI in technical duplicates across two independent experiments. Successful entry was monitored by luciferase activity in cell lysate 48 hours after inoculation (**Figure 4b, Supplementary Fig. 5a**). As previously reported, the D614G mutation significantly enhanced viral entry over the original Spike containing D614. Two mutations enhanced viral entry even further in conjunction with the D614G mutation: the 69-70 deletion (FDR<0.001) and the Q52R point mutation (FDR=0.0252). One mutation, the 144 deletion, appeared to negatively impact viral entry when combined with D614G alone. Nevertheless, this defect must be compensated for by the additional mutations as the Spike protein with all 8 mutations found in B.1.525 showed the highest amount of viral entry relative to each tested mutation alone. Together, these results suggest that the Spike mutations in the B.1.525 lineage enhance cell entry, consistent with the phenotypic impact of Spike mutations in other lineages of concern ^48,49^.

### Spike protein mutations in the B.1.525 lineage reduce antibody neutralization after natural infection

One major concern for any new SARS-CoV-2 Spike mutation is its potential impact on antibody-mediated neutralization after vaccination or natural infection. To examine the susceptibility of viruses with B.1.525 lineage Spike protein to neutralizing antibodies, HIV-1 luciferase reporter viruses were pseudotyped with a subset of the mutations tested above: either the original D614 Spike protein, Spike protein containing the D614G mutation, or Spike protein containing the 8 mutations that characterize the B.1.525 lineage. Viruses were normalized by p24 concentration and incubated with sera isolated from individuals inoculated with two doses of the BNT162b2 vaccine (Pfizer, n = 4), individuals inoculated with two doses of the mRNA-1273 vaccine (Moderna, n = 4), or convalescent individuals recovered from COVID-19 infection (n = 9) (**Supplemental Table 2)**. Serum from vaccinated individuals was collected roughly two weeks after the second vaccine dose had been administered. Convalescent serum had been collected from residents of Oyo state, Nigeria prior to the emergence of the B.1.525 lineage (May 18^th^ 2020 and June 27^th^ 2020) at least 5 days, but no later than 5 weeks, after diagnosis with SARS-CoV-2 infection. Antibodies against Spike RBD and NTD (due to the accumulation of mutations in this region) were quantified by ELISA in all serum specimens to confirm the presence of an immune response prior to neutralization testing (**Supplementary Fig. 5b**). On average, serum from vaccinated individuals carried higher antibody titers against both RBD and NTD than convalescent serum. Concentrations of RBD and NTD antibodies in all individuals showed a very strong correlation with each other (Spearman rho = 0.93; **Supplementary Fig. 5c**). After incubation in different dilutions of serum, pseudotyped viruses expressing each Spike mutation were used to challenge HeLa-ACE2 cells with infection measured by luciferase activity in cell lysates after 48 hours. The half maximal effective concentration (EC_50_) was subsequently extrapolated from each dilution curve for both Spike RBD and NTD antibodies (**Supplementary Fig. 6a and 6b**).

Serum from individuals fully vaccinated with either mRNA vaccine was highly protective against all Spike variants tested, with no significant differences observed between neutralization of viruses expressing the original D614 Spike, G614 Spike, or B.1.525 Spike (**Figure 4c**). Convalescent sera similarly demonstrated nearly complete neutralization of viruses carrying D614 and G614 Spike variants when compared to controls without serum treatment. However, the ability of convalescent sera to neutralize B.1.525 Spike RBD (median EC_50_ = 0.17 µg/ml) and NTD (median EC_50_ = 1.26 µg/ml) was significantly restricted compared to either D614 (RBD median EC_50_ = 0.04 µg/ml, NTD median EC_50_ = 0.03 µg/ml) or G614 Spike (RBD median EC_50_ = 0.02 µg/ml, NTD median EC_50_ = 0.07 µg/ml) (**Figure 4c**). Overall Spike RBD and NTD antibody levels in each specimen correlated with the neutralization activity of D614 Spike (**Supplementary Fig. 5d**), suggesting that increased antibody titer following vaccination may be one correlate of improved protection. However, these results also suggest that the B.1.525 Spike protein may confer some protection against antibody-mediated neutralization following natural infection with a different lineage.

## DISCUSSION

These results demonstrate the emergence and subsequent dominance of two distinct viral lineages in Oyo state, Nigeria: the B.1.1.7 Alpha ‘variant of concern’ and the B.1.525 Eta ‘variant of interest’. As variants with documented evidence of enhanced infectivity, transmissibility, and/or immune evasion continue to emerge and expand across the globe, updated guidance is needed for improved diagnostic and prevention strategies in ‘variant dominant’ countries and locales. In addition, documented substantial expansion of B.1.525, coupled with the evidence of increased infectivity *in vitro* and enhanced immune evasion after natural infection, suggest that B.1.525 should be elevated to a ‘variant of concern’ from its current designation as a ‘variant of interest’. More broadly, this work demonstrates the critical need for international cooperation in infectious disease surveillance in undersampled regions for the monitoring and detection of new pathogenic threats.

The B.1.1.7 lineage, which was first reported on December 21, 2020 in the U.K. and first sampled on September 20, 2020, appears to have become dominant within Oyo state and throughout the other surveilled states in Nigeria as of early January 2021. While this lineage has not been found to influence efficacy of neutralizing antibodies produced by vaccination, it is considered a variant of concern as it expanded rapidly in multiple places across the globe, was found to be more infectious *in vitro*, has evidence of increased transmissiblity, and can interfere with diagnostic tests probing over the 69-70 deletion in the S protein ^20,50-52^. Phylogenetic analysis suggests a single or multiple, closely linked introductions of the B.1.1.7 virus lineage into Nigeria in the fall of 2020.

Compared to the B.1.1.7 lineage, many fewer representative cases of the B.1.525 lineage have been reported to date, although it has also expanded globally. The highest proportion of B.1.525 is observed in West Africa, including Nigeria, with phylogenetic analyses of the available sequences consistent with this region being the source of other cases observed elsewhere globally. That being said, we cannot conclude that the B.1.525 lineage originated in Nigeria due to low sampling and the observation that two sequences from Spain root outside of the node with the highest statistical support corresponding to TMRCA for the B.1.525 lineage in Nigeria. Importantly, this lineage carries several Spike mutations that have been previously linked to transmissibility and decreased vaccine efficacy, leading to it being designated a ‘variant of interest’ shortly after its description. Not only does B.1.525 share two Spike deletions with B.1.1.7 (69-70del and 144del), but this lineage also contains the Spike E484K mutation, which prior studies had suggested could impact immune recognition and vaccine efficacy ^53-58^. Similar to B.1.1.7, phylogenetic analysis of the B.1.525 lineage in Nigeria suggests either a single or multiple, closely linked introductions of the virus into Nigeria in the fall of 2020.

While these analyses suggest both lineages arose from either single or multiple, closely linked introduction events, overall lack of sampling across this region remains a significant confounder and more data will be needed to further test these hypotheses. Comparing the estimated TMRCA for both lineages with the daily incidence reported by the Nigerian Centers for Disease Control (**Figure 5**), we note that the possible introduction or appearance of these lineages coincides with some of the lowest daily incidence numbers since the beginning of the pandemic. This observation suggests the possibility that both lineages benefited from founder effects that boosted overall prevalence of each as case numbers rose. Less than a month prior to the estimated TMRCA for these lineages, Nigeria reopened international airports to regular air traffic (September 5, 2020). This step was accompanied by strong measures to prevent SARS-CoV-2 introduction to the country, including requiring a negative qRT-PCR test within 96 hours of boarding an in-bound flight, screening for COVID-19 symptoms prior to boarding the flight, self-quarantine for 7 days after arrival in Nigeria, and a mandatory second qRT-PCR test on day 7 of arrival. Despite these rigorous public health measures, these data are consistent with a model wherein these lineages could have been seeded from international travel followed by local expansion.

**Figure 5.**
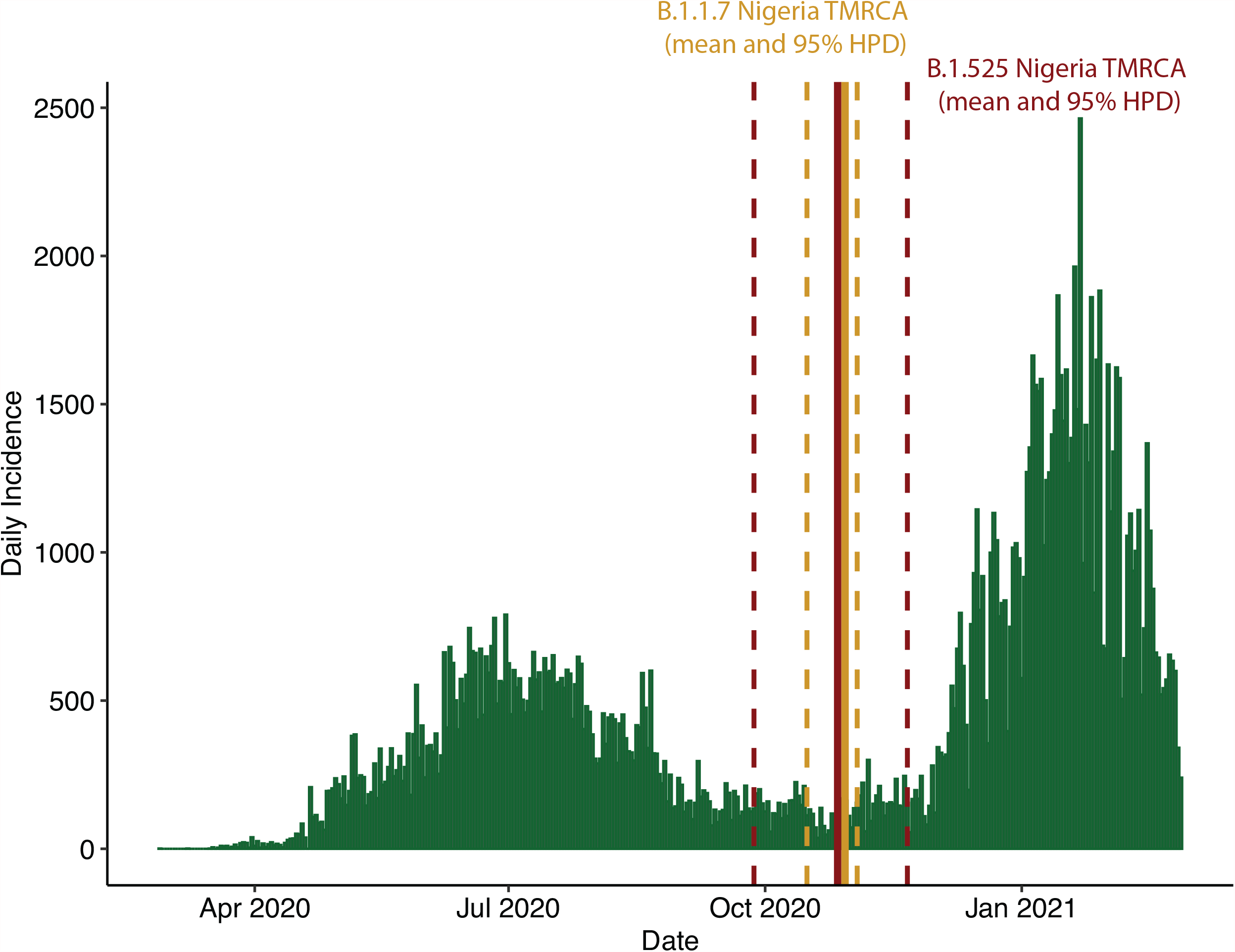
Daily SARS-CoV2 Incidence in Nigeria. Confirmed new cases in Nigeria obtained from Johns Hopkins University Coronavirus resource center (https://coronavirus.jhu.edu/). The TMRCA (solid line) and 95% High Probability Density (HPD) (dashed lines) in Nigeria of B.1.1.7 and B.1.525 lineages estimated using Bayesian methods is indicated.

While founder-effects may have contributed to the predominance of these strains in Nigeria following their introduction during a nadir in case counts, cell culture entry analyses conducted using Spike-pseudotyped virus particles suggest that the B.1.1.7 and B.1.525 Spike proteins may also enhance viral entry and confer a selective advantage. Previous reports have described incremental improvement in SARS-CoV-2 cell entry through Spike protein mutations in lineages that subsequently expanded internationally. The D614G mutation in Spike is now nearly fixed across the global population and defines the predominant B.1 lineage from which the above two lineages derivate. This mutation was found to improve viral entry into airway epithelial cells and confer both increased transmissibility and increased fitness^10-15^. Subsequently, multiple further variations in Spike, including the 69-70del in the B.1.1.7 lineage, have been described that improve viral entry even further over the G614 background ^19,41,43,49,51,57,59^. Intriguingly, the 69-70 deletion, as well as Q52R to a lesser degree, significantly increased Spike mediated cell entry in our assays despite being located far from the ACE2 binding site (**Figure 4a and 4b**). A deeper examination of this region is needed to understand possible effects of these mutations on receptor binding or the mechanics of viral entry.

In addition to increased efficiency of viral entry, we also observed decreased antibody neutralization of viruses carrying B.1.525 Spike proteins in convalescent serum from individuals living in Oyo state who had previously had COVID-19 infections. Other SARS-CoV-2 lineages have also been shown to have increased potential for immune evasion in convalescent sera^58^, including the P.1 lineage, which shares a Spike E484K mutation with B.1.525 ^22^. The E484K mutation has been shown to arise spontaneously in viruses passaged in convalescent serum ^60^, further supporting its role in potentially protecting the virus from antibody responses against other non-variant Spike proteins ^61,62^. Moreover, most of the S gene mutations acquired by B.1.525 are located in the NTD of the Spike protein, possibly affecting a known antigenic site ^63,64^. This observation suggests likely additional effects of this set of mutations on natural immunity, specifically on antibodies directed at the NTD of the protein^65^. The decreased neutralization levels observed when comparing NTD antibody concentrations in this study were more significant than when using RBD concentrations (**Figure 4c)**. Although our sample size is limited, this observation might suggest a further impairment in the Spike recognition by NTD directed antibodies due to the accumulation of mutations in this region.

The successful introduction and/or spread of these lineages in a period of very low incidence, together with their possible fitness advantages, could have facilitated the coincident rise and dominance of these lineages in Nigeria. The ability of the B.1.525 lineage to expand alongside B.1.1.7, the observed increase in infectivity of B.1.525 Spike containing viruses, and the decrease in antibody neutralization of B.1.5252 Spike virus by convalescent serum all suggest it is a variant of concern. More recently, as of early June 2021, B.1.525 appears to have outcompeted B.1.1.7 in Nigeria and has become the dominant lineage in the country (https://covariants.org/, https://coronavirus3d.org/). Although this observation is derived from a very limited number of public sequences, it is suggestive of a higher infectivity rate of B.1.525 over the already highly infective B.1.1.7. Therefore, it is imperative to monitor this lineage and better understand its real-world impact in Nigeria and the rest of West Africa, especially as vaccination programs in the region get underway. Additional sequencing and surveillance across undersampled regions, and elucidation of their biological and clinical significance, will be required to better understand and cope with emerging variants on a global scale.

## METHODS

### Viral RNA extraction

Viral RNA was extracted from clinical specimens utilizing the QIAamp Viral RNA Minikit (Qiagen, cat. no. 52906). Clinical testing for SARS-CoV-2 presence was performed by quantitative reverse transcription and PCR (qRT-PCR) with the CDC 2019-nCoV RT-PCR Diagnostic Panel utilizing the N1 probe in SARS-CoV-2 and RNase P probes for sample quality control as previously described (IDT, cat. no. 10006713). All specimens that failed to amplify the RNase P housekeeping gene were excluded from this study. All specimens with an N1 probe cycle threshold (Ct) less than or equal to 35 were considered positive and included in this study. qRT-PCR was repeated in a random selection of specimens to validate Ct values obtained by the clinical diagnostic laboratory. Ct values from the N1 probes were used in all subsequent analyses.

### cDNA synthesis and viral genome amplification

cDNA synthesis was performed with SuperScript IV First Strand Synthesis Kit (ThermoFisher, cat. no. 18091050) using 11 μl of extracted viral nucleic acids and random hexamers according to manufacturer’s specifications. Direct amplification of the viral genome cDNA was performed in multiplexed PCR reactions to generate ∼400 bp amplicons tiled across the genome. The multiplex primer set, comprised of two non-overlapping primer pools, was created using Primal Scheme and provided by the Artic Network (version 3 release). PCR amplification was carried out using Q5 Hot Start HF Taq Polymerase (NEB, cat. no. M0493L) with 5 μl of cDNA in a 25 μl reaction volume. A two-step PCR program was used with an initial step of 98 °C for 30 s, then 35 cycles of 98 °C for 15 s followed by five minutes at 64 °C. Separate reactions were carried out for each primer pool and validated by agarose gel electrophoresis alongside negative controls. Each reaction set included positive and negative amplification controls and was performed in a space physically separated for pre- and post-PCR processing steps to reduce contamination. Amplicon sets for each genome were pooled prior to sequencing library preparation.

### Sequencing library preparation, Illumina sequencing, and genome assembly

Sequencing library preparation of genome amplicon pools was performed using the SeqWell plexWell 384 kit per manufacturer’s instructions. Pooled libraries of up to 96 genomes were sequenced on the Illumina MiSeq using the V2 500 cycle kit. Sequencing reads were trimmed to remove adapters and low-quality sequences using Trimmomatic v0.36. Trimmed reads were aligned to the reference genome sequence of SARS-CoV-2 (accession MN908947.3) using bwa v0.7.15. Pileups were generated from the alignment using samtools v1.9 and consensus sequence determined using iVar v1.2.2 ^66^ with a minimum depth of 10, a minimum base quality score of 20, and a consensus frequency threshold of 0 (i.e., majority base as the consensus).

### Phylogenetic analysis

Genome sequences were aligned using MAFFT v7.453 software ^67^ and manually edited using MEGAX v10.1.8^68^. All Maximum Likelihood (ML) phylogenies were inferred with IQ-Tree v2.0.5^69^ using its ModelFinder function^70^ before each analysis to estimate the nucleotide substitution model best-fitted for each dataset by means of Bayesian information criterion (BIC). We assessed the tree topology for each phylogeny both with the Shimodaira–Hasegawa approximate likelihood-ratio test (SH-aLRT) ^71^ and with ultrafast bootstrap (UFboot) ^72^ with 1000 replicates each. TreeTime v0.7.6 ^73^ was used for the assessment of root-to-tip correlation, the estimation of time scaled phylogenies and ancestral reconstruction of most likely sequences of internal nodes of the tree and transitions between geographical locations along branches. TreeTime was run using an autocorrelated molecular clock under a skyline coalescent tree prior. We used the sampling dates of the sequences to estimate the evolutionary rates and determine the best rooting of the tree using root-to-tip regression with least-squares method.

Bayesian time-scaled phylogenetic analyses were performed for the phylogenies of B.1.1.7 and B.1.525 lineages separately using a smaller set (for computational time reasons) of 500 randomly sampled global sequences from GISAID for B.1.1.7 and all the available sequences for B.1.525. Due to the small number of B.1.525 sequences available, we included B.1 sequences from our Nigerian dataset as well as 10 randomly selected B.1 sequences from GISAID to help root the phylogenies. We used BEAST v2.5.2 ^74^ to estimate the date and location of the most recent common ancestors (MRCA) as well as to estimate the rate of evolution of the virus. BEAST priors were introduced with BEAUTI v2.5.2 including an uncorrelated relaxed molecular clock model with a lognormal distribution of the evolutionary rate, previous estimated evolutionary rates (8×10-4) as the prior for the mean, and a standard deviation of 0.1 after optimization with preliminary runs. We assumed a GTR substitution model with invariant sites, as the best-fitted model obtained with ModelFinder, and a Coalescence Bayesian Skyline to model the population size changes through time. The posterior evolutionary rate was estimated to be 5.9×10-4 for the B.1.1.7 inference and 7.9×10-4 for the B.1.525 analysis (that included B.1 lineages). All the analyses indicated support for a relaxed clock as the average rate coefficient of variation was 0.2 for B.1.1.7 and 0.6 for B.1.525, which indicates a degree of rate autocorrelation among adjacent branches in the tree. Markov chain Monte Carlo (MCMC) runs of at least 100 million states with sampling every 5,000 steps were computed. The convergence of MCMC chains was monitored using Tracer v.1.7.1, ensuring that the effective sample size (ESS) values were greater than 200 for each parameter estimated.

### Cell lines

To produce HeLa cells (obtained from AIDS reagents) overexpressing Ace2, HEK-293T cells were transfected with psPAX2, pCMV-VSVG, and pRRL.sin.cPPT.SFFV/Ace2.IRES-puro.WPRE (a gift from Caroline Goujon; Addgene plasmid # 145839; http://n2t.net/addgene:145839; RRID:Addgene_145839). HeLa cells were then transduced with the lentiviral particles and puromycin (Invivogen) selected as in ^75^. Overexpression was validated by Western blotting.

### Generation of SARS-CoV-2 pseudovirus

HEK-293T cells were transfected with a 3:2 ratio of NL4-3-nanoluc delta env plasmid (gift from Thomas Hope) and pCAGGS-Spike Sars-CoV2 plasmid (and introduced mutations) respectively. (The following reagent was produced under HHSN272201400008C and obtained through BEI Resources, NIAID, NIH: Vector pCAGGS Containing the SARS-Related Coronavirus 2, Wuhan-Hu-1 Spike Glycoprotein Receptor Binding Domain (RBD), NR-52309). Culture media was changed 16 hours post transfection and viral particles were harvested at 48 hours. Viral particles were concentrated using a 20% sucrose gradient with overnight centrifugation at 5,600 rcf at 4°C and then resuspended into fresh media at a 500x concentration. The concentrated Spike pseudoviruses were quantified using HIV-1 Gag p24 Quantikine ELISA Kit (R&D Systems), and the infectivity was determined using Hela-Ace2 cells and the Nano-Glo® Dual-Luciferase® Reporter Assay (Promega).

### Synthesis of B.1.525 lineage mutations

The introduction of the B.1.525 lineage mutations into pCAGS-Spike Sars-CoV2 plasmids (BEI) was performed by restriction digest of the plasmid, followed by NEBuilder HiFi DNA assembly (New England Biolabs) with either amplified by PCR products or synthesized DNA fragments (Genewiz). The full modified sequences were verified and confirmed by Sanger sequencing (Supplementary Methods).

### Antibody neutralization assay

HeLa-Ace2 cells were plated in 96-well black wall plates at 10^5^ cells per well and incubated at 37°C, with 5% CO_2_ for 24 hours. Media was then removed, and serum was added using serial dilutions. Pseudotyped virus (pCAGGS-Spike Sars-CoV2 and mutants; NL4-3-nanoluc delta env) was added to each well, and cells were returned to the incubator. After a 48-hour incubation, reporter gene expression was determined with Nano-Glo® Dual-Luciferase® Reporter Assay (Promega) following the vendor’s protocol using a luminometer (Cytation3, Biotek). We estimated the amount of RBD-binding and NTD-binding antibodies at half maximal effective concentration (EC_50_) for each serum sample by fitting a four-parameter log-logistic function to each dilution curve for both RBD and NTD using the drc package in R v4.0.3. For this estimation, we used the luminescence values measured in relative light units (RLU) normalized both by the values obtained with the negative controls without virus and sera and the positive controls with each corresponding virus and no sera added (**Supplementary Fig. 6c**).

### RBD- and NTD-binding by ELISA

Ni-NTA HisSorb 96-well plates (Qiagen) were coated with 50ul of either His-tagged RBD or His-tagged NTD (Acro Biosystems) at a concentration of 2ug/ml overnight at 4°C; all subsequent steps were performed at room temperature. Plates were blocked with 3% milk; plasma was diluted 1:500 in PBS for RBD ELISA and 1:1500 for NTD ELISA prior to adding 100ul per well for 2 hours. Plates were washed with PBS-T 3 times, followed by incubation with secondary anti-human IgG Fc HRP (Southern Biotech) at 1:4000 for 1hr. Plates were washed 3 additional times with PBS-T, followed by addition of TMB substrate for 10min. The reaction was stopped with the addition of 3M HCl; and plates were read at an OD 450. Anti-RBD antibody CR3022 (Creative Biolabs) was used to make a standard curve with 1:2 dilutions in the range of 0.5ug/ml to 0.03ug/ml, utilizing a 4-parameter fit to interpolate sample concentrations. Anti-NTD antibody SPD-M121 (Acro Biosystems) was used to make a standard curve with 1:2 dilutions in the range of 0.5ug/ml to 0.03ug/ml, utilizing a 4-parameter fit to interpolate sample concentrations.

### Structural modeling of B.1.525 S protein

A model of the B.1.525 NTD domain, with the Q52R and A67V mutations and H69, V70 and Y144 deletions was built using SwissModel ^76^ using a PDB coordinate of the Fab C25 complex with the NTD of SARS-CoV-2 (PDB 7m8j) ^77^ as a template. The complex was minimized and binding energy was estimated using the EvoEF program, part of the EvoDesign pipeline ^78,79^.

### Statistical Analyses

All statistical analyses were performed in in R v 4.0.3. Statistical analysis of cell entry and neutralization EC_50_ were performed fitting Linear Mixed-Effects Models using lme4 package. For cell entry we tested for significant differences in cell entry between mutants with increasing concentrations of psudotyped viruses including the interaction between mutant and p24 concentration in the model and accounting for experiment and plate effects. For neutralization EC_50_ differences we included in the model the different groups of sera, each mutant, as well as the interaction of both parameters, with the addition of the possible effect of within-donor sera correlation. Afterwards we performed every possible contrast within each model and used FDR to assess significance while controlling for multiple comparisons. A linear model including patient sex and age as confounders was fitted to test for differences in Ct value between lineages.

## Data Availability

The final 74 complete SARS-CoV-2 genomes were deposited in the publicly available GISAID database (Supplemental Table 1)

## ACKNOWLEDGEMENTS

This research was supported in part through the computational resources and staff contributions provided for the Quest high performance computing facility at Northwestern University, which is jointly supported by the Office of the Provost, the Office for Research, and Northwestern University Information Technology. Genome sequencing and analysis was supported by resources and staff at the Center for Pathogen Genomics and Microbial Evolution in the Northwestern University Institute for Global Health. Funding for this work was provided by: a Northwestern Institute for Global Health Catalyzer Research Fund award (R.L.R. and O.M.A.); a Dixon Translational Research Grant made possible by the generous support of the Dixon Family Foundation (E.A.O. and J.F.H.); a COVID-19 Supplemental Research award from the Northwestern Center for Advanced Technologies (NUCATS - J.F.H.); a CTSA supplement to NCATS UL1 TR002389 (J.F.H., E.A.O., R.L.R.); a supplement to the Northwestern University Cancer Center P30 CA060553 (J.F.H.); the Gilead Sciences Research Scholars Program in HIV (J.F.H.); the NIH-supported Third Coast CFAR P30 AI117943 (R.L.R., J.F.H.); NIH grant K22 AI136691 (J.F.H.); NIH grant U19 AI135964 (E.A.O.); NIH grant D43 TW009608 (B.O.T., O.M.A.); NIH Fogarty International Center award D43TW009608 (B.O.T.); NIH grant R35 GM118187 (A.G.) and NIH-NIAID Center for Structural Genomics of Infectious Diseases award HHSN272201700060C (A.G.); and through a generous contribution from the Walder Foundation Foundation’s Chicago Coronavirus Assessment Network (Chicago CAN) Initiative (J.F.H., E.A.O., R.L.R.) and Walder Foundation grant numbers SCI16(J.R.S) and 21-00147(J.R.S.). The authors also acknowledge the Nigeria Center for Disease Control (NCDC) for providing leadership and infrastructure that facilitated collection of samples included in this study. The funding sources had no role in the study design, data collection, analysis, interpretation, or writing of the report.

## AUTHOR CONTRIBUTIONS

Conceptualization: R.L.R., L.M.S., J.F.H., E.A.O., B.O.T. Investigation: O.M.A., A.A.F., E.C.O., J.A.A., L.M.S., T.J.D., J.Z., P.P.B., M.K.A., A.G., J.R.S., J.I.M., R.L.R., J.F.H., E.A.O. Resources: O.M.A., J.F.H., E.A.O., J.R.S., J.I.M. Formal analysis: R.L.R., E.A.O. Supervision: J.F.H., E.A.O. Funding acquisition: J.F.H., E.A.O., R.L.R., B.O.T., J.R.S Writing: R.L.R., L.M.S., J.F.H., E.A.O., B.O.T., O.M.A., A.G., J.R.S., J.I.M.

## FIGURE LEGENDS

**Supplementary Figure 1.**
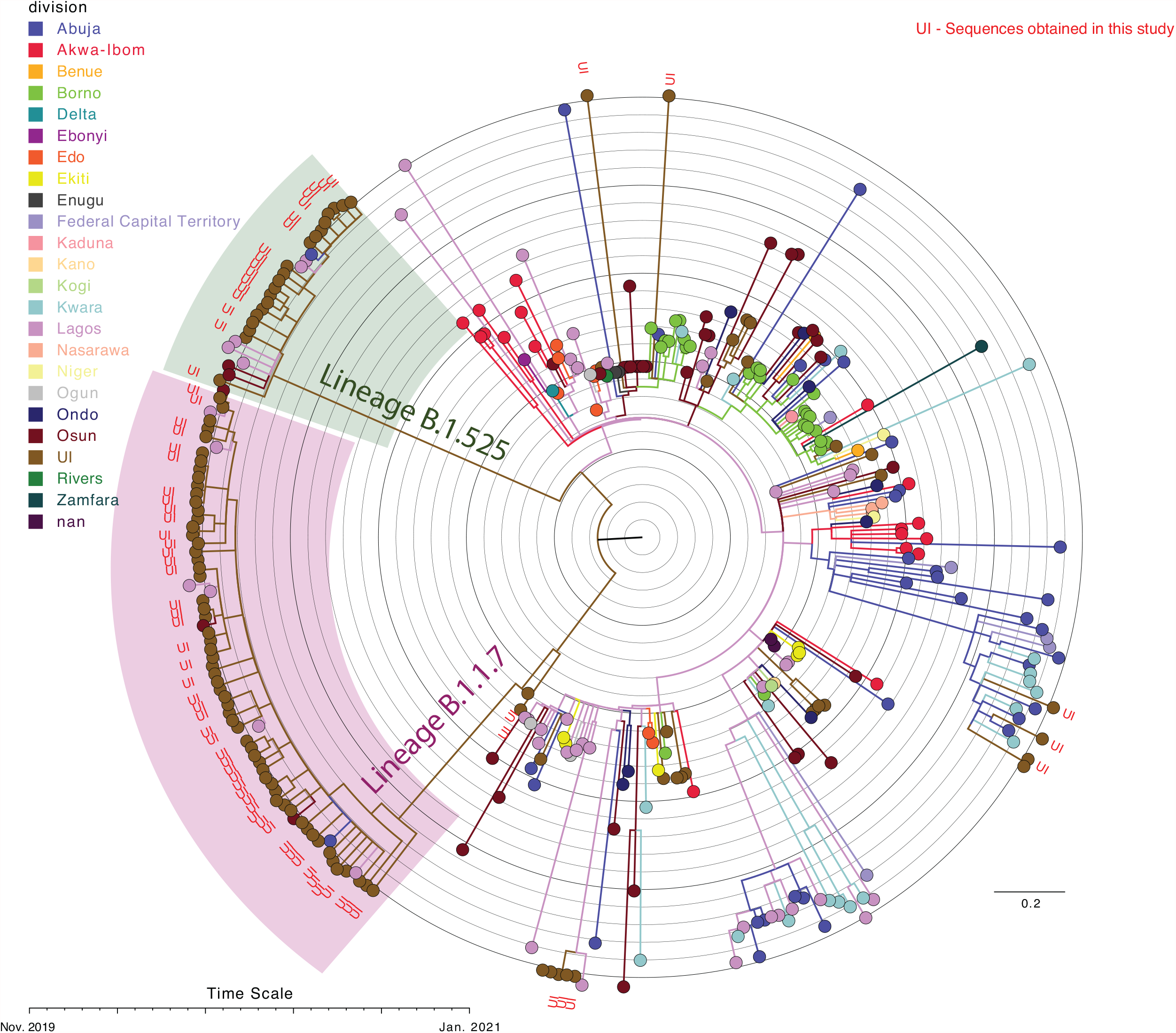
Phylogenetic analysis of Oyo state isolates compared to all available Nigerian sequences. ML phylogenetic temporal reconstruction of full genome sequences from Nigeria, including the sequences from this study and all sequences available from Nigeria in GISAID as of February 14^th^, 2021. Clades corresponding to B.1.1.7 and B.1.525 lineages are indicated. Branches and tips are colored by state; labels corresponding to sequences obtained in this study are colored in red.

**Supplementary Figure 2.**
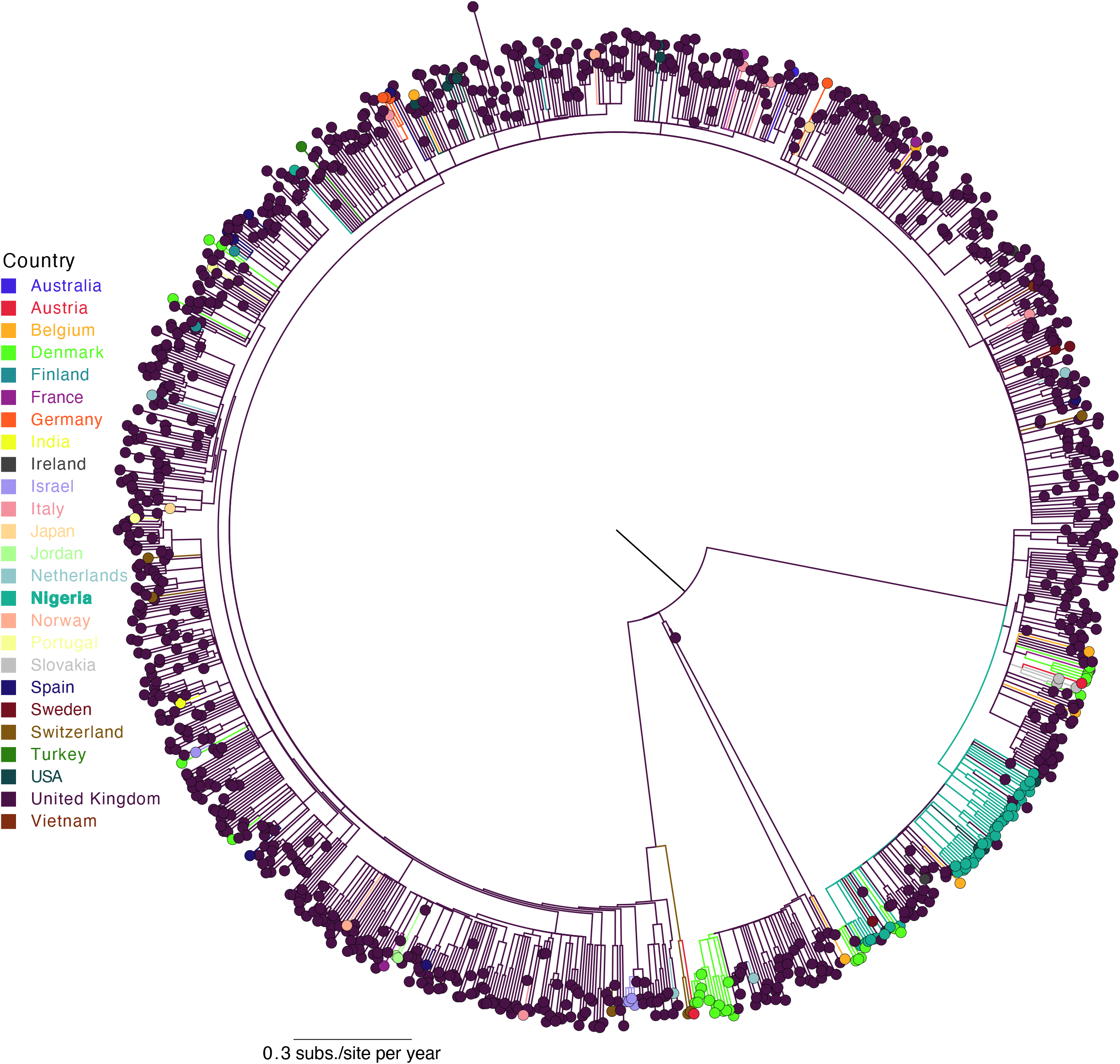
Phylogenetic analysis of Oyo state B.1.1.7 sequences compared to global B.1.1.7 sequences. ML phylogenetic temporal reconstruction of full genome B.1.1.7 sequences obtained in this study and 4000 randomly sampled B..1.1.7 global sequences from GISAID as of February 14^th^, 2021. Branches and tips are colored by country.

**Supplementary Figure 3.**
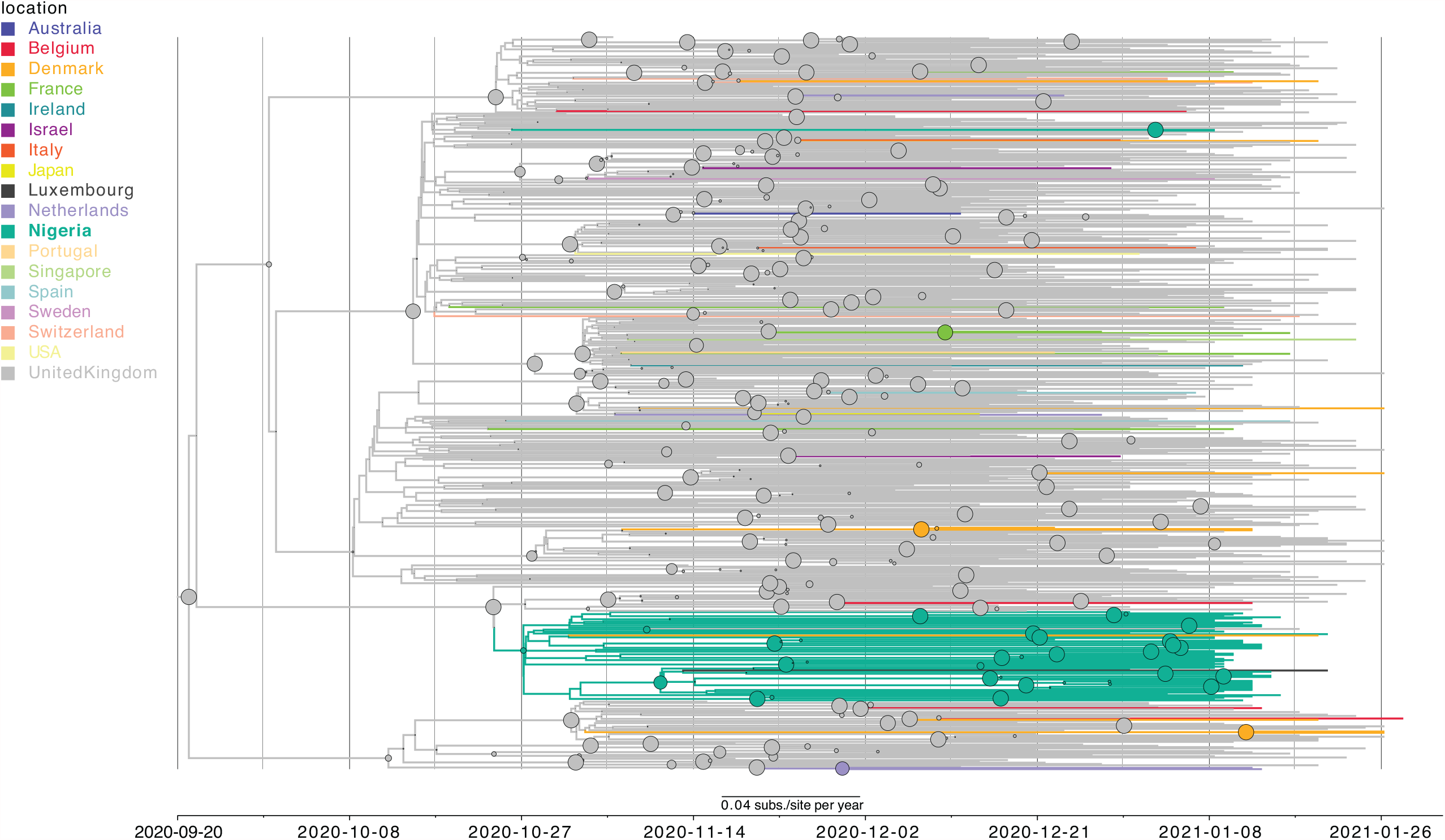
Phylodynamic tree of Nigeria B.1.1.7 sequences compared to global B.1.1.7 sequences. Maximum clade credibility tree where branch colors represent the most probable geographical location of their descendent node inferred through Bayesian reconstruction of the ancestral state. All full genome B.1.1.7 sequences from Nigeria and 500 randomly sampled B.1.1.7 global sequences from GISAID as of February 14th, 2021 were included in the analysis. The width of the node circles represents their posterior probability.

**Supplementary Figure 4.**
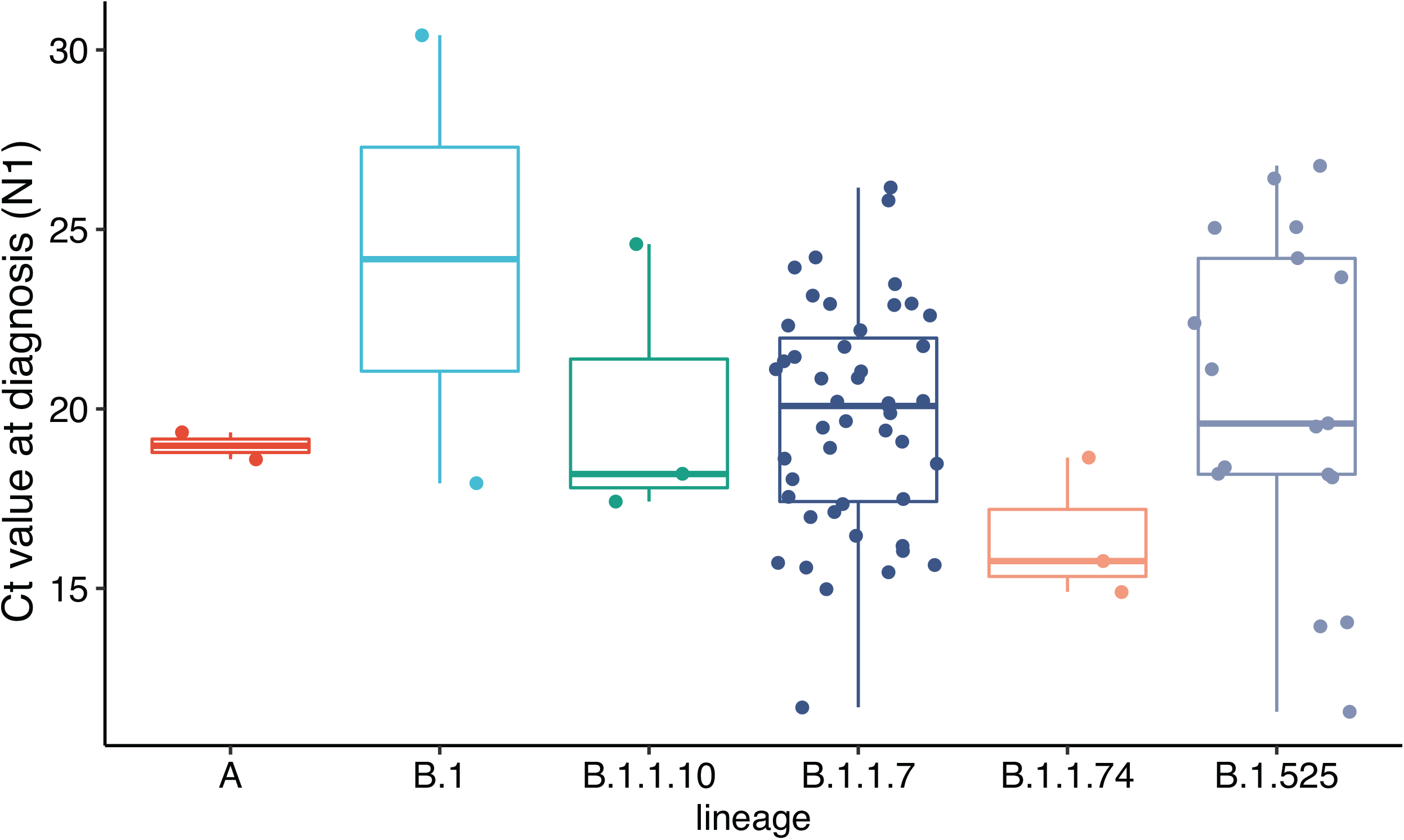
PCR Cycle threshold (Ct) values of patient diagnostic samples grouped by pangolin lineage assignment. Ct values for the N1 probe set reported at the time of diagnosis were compared between lineages. A linear model was fitted to test for differences in Ct value between lineages. All possible contrasts within the model were performed and corrected for multiple comparisons using Tukey’s test. No corrected p-values were found to be statistically significant using 0.05 cut-off. Tukey’s box and whisker plots; box limits: interquartile range (IQR); middle line: median

**Supplementary Figure 5.**
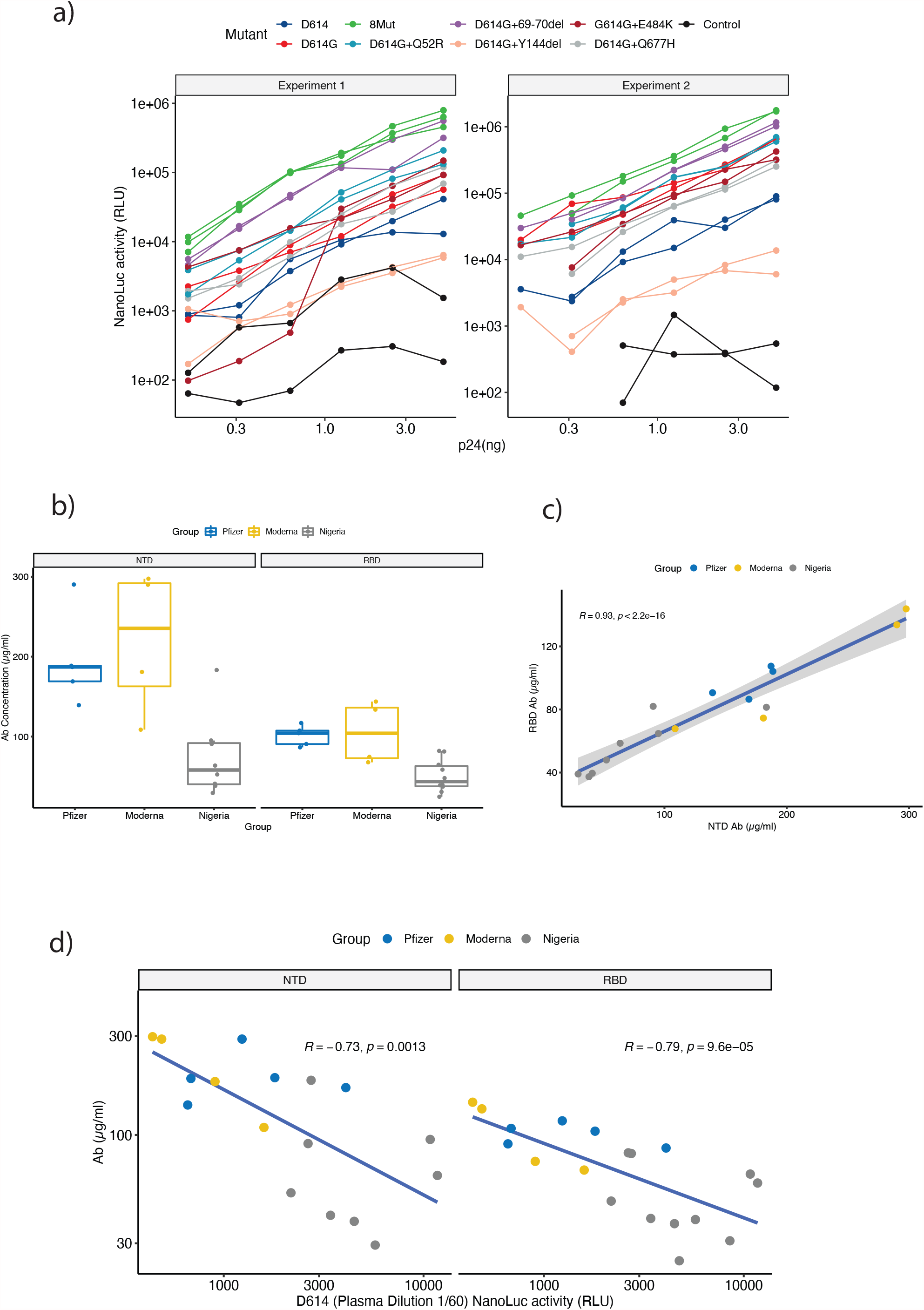
Cell entry of pseudotyped viruses with B.1.525 Spike mutations and Spike antibody concentrations in the tested sera. a) Nanoluc activity measured in relative light units (RLU) of the mutants tested, including D614G and D614. Lines colored by mutant represent replicates for each mutant in the two independent experiments performed. Dots are nanoluc activity values per reporter virus concentration. b) Spike NTD-binding and RBD-binding antibody concentrations calculated for each of the sera used in the neutralization experiments. Tukey’s box and whisker plots; box limits: interquartile range (IQR); middle line: median. c) Correlation between RBD and NTD antibodies in each serum. d) Correlation between RBD and NTD antibodies and Nanoluc activity of D614 mutant at the lowest dilution used in the experiments (1:60). A very significant negative correlation between antibody concentration and reporter virus is found for both NTD and RBD indicating their effective neutralization of D614 mutant. For c and d dots are colored by the sera group, line represents linear regression and grey area is the regression confidence interval. Spearman rho and p-value are shown.

**Supplementary Figure 6.**
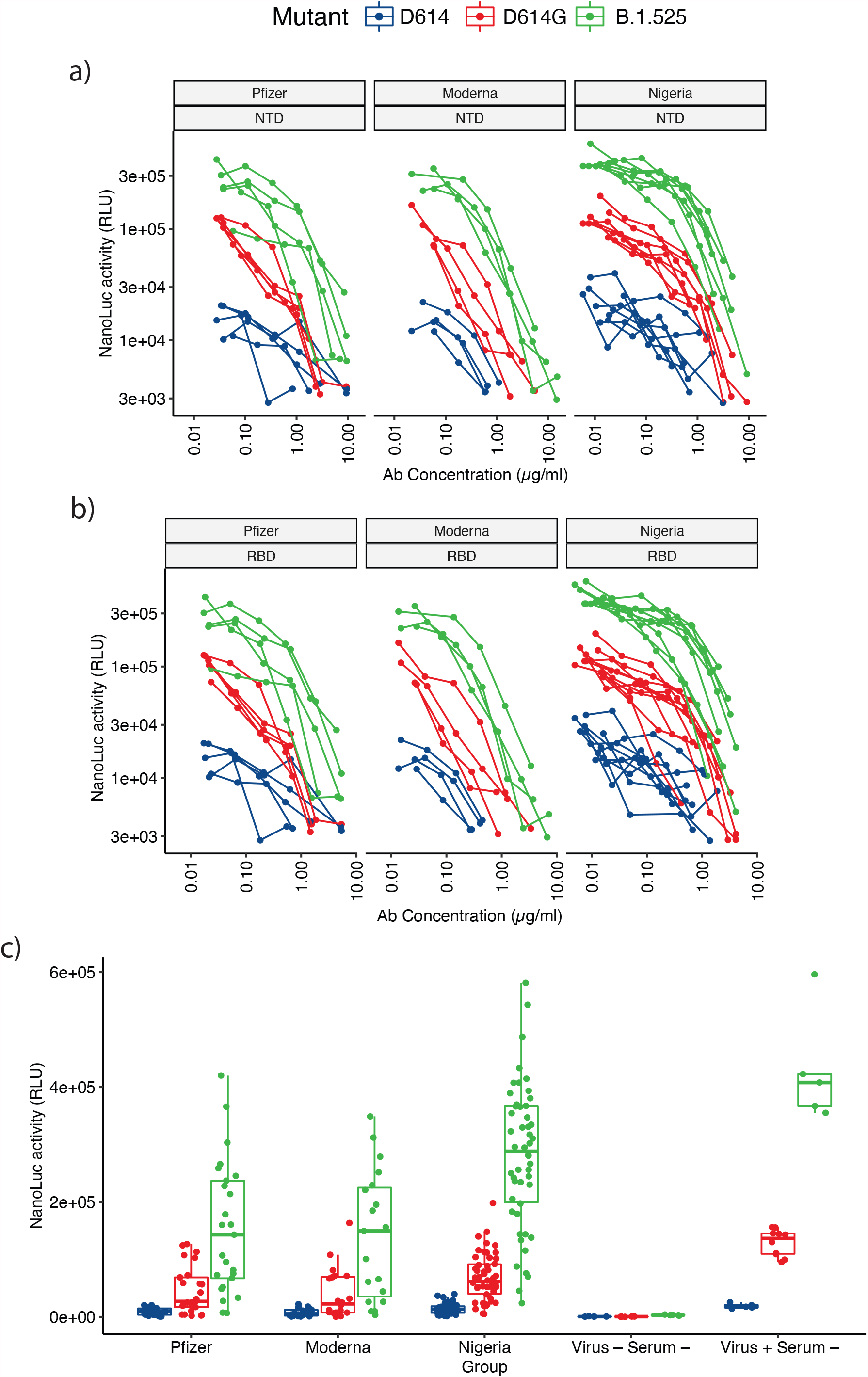
Dilution curves for both NTD-binding and RBD-binding antibodies and all nanoluc activity values. a) Nanoluc activity values per NTD-binding antibody concentration. b) Nanoluc activity values per RBD-binding antibody concentration. For both a) and b) lines represent each serum and dots are the nanoluc activity values per each serum dilution. Lines are colored by the mutant tested for every serum. c) Boxplot representing all nanoluc activity luminescence RLU values obtained in the neutralization experiments including the controls that were used to normalize these luminescence values for EC_50_ estimation. Boxes are colored by mutant and dots represent each luminescence value. Positive controls included virus without serum (Virus+ Serum-) and negative controls did not include either virus or sera (Virus-Serum-).

