## Supplementary Table 1 for "Coincident rapid expansion of two SARS-CoV-2 lineages with enhanced infectivity in Nigeria"

| GISAID Virus ID | GISAID Accession | Isolation Date | Pango lineage |
| --- | --- | --- | --- |
| hCoV-19/Nigeria/BCVL-18292/2021 | EPI_ISL_985102 | 2021-01-02 | B.1.1.7 |
| hCoV-19/Nigeria/BCVL-18921/2021 | EPI_ISL_985124 | 2021-01-05 | B.1.525 |
| hCoV-19/Nigeria/BCVL-18912/2021 | EPI_ISL_985238 | 2021-01-05 | A.27 |
| hCoV-19/Nigeria/BCVL-18900/2021 | EPI_ISL_985123 | 2021-01-06 | B.1.525 |
| hCoV-19/Nigeria/BCVL-18568/2021 | EPI_ISL_985058 | 2021-01-07 | B.1.1.7 |
| hCoV-19/Nigeria/BCVL-18510/2021 | EPI_ISL_985091 | 2021-01-07 | B.1.1.7 |
| hCoV-19/Nigeria/BCVL-18521/2021 | EPI_ISL_985092 | 2021-01-07 | B.1.1.7 |
| hCoV-19/Nigeria/BCVL-18523/2021 | EPI_ISL_985093 | 2021-01-07 | B.1.1.7 |
| hCoV-19/Nigeria/BCVL-18543/2021 | EPI_ISL_985094 | 2021-01-07 | B.1.1.7 |
| hCoV-19/Nigeria/BCVL-18302/2021 | EPI_ISL_985095 | 2021-01-07 | B.1.1.7 |
| hCoV-19/Nigeria/BCVL-18303/2021 | EPI_ISL_985096 | 2021-01-07 | B.1.1.7 |
| hCoV-19/Nigeria/BCVL-18308/2021 | EPI_ISL_985097 | 2021-01-07 | B.1.1.7 |
| hCoV-19/Nigeria/BCVL-18598/2021 | EPI_ISL_985098 | 2021-01-07 | B.1.1.7 |
| hCoV-19/Nigeria/BCVL-18611/2021 | EPI_ISL_985099 | 2021-01-07 | B.1.1.7 |
| hCoV-19/Nigeria/BCVL-18631/2021 | EPI_ISL_985100 | 2021-01-07 | B.1.1.7 |
| hCoV-19/Nigeria/BCVL-18632/2021 | EPI_ISL_985101 | 2021-01-07 | B.1.1.7 |
| hCoV-19/Nigeria/BCVL-18560/2021 | EPI_ISL_985103 | 2021-01-07 | B.1.1.7 |
| hCoV-19/Nigeria/BCVL-18639/2021 | EPI_ISL_985104 | 2021-01-07 | B.1.1.7 |
| hCoV-19/Nigeria/BCVL-18562/2021 | EPI_ISL_985106 | 2021-01-07 | B.1.1.318 |
| hCoV-19/Nigeria/BCVL-18579/2021 | EPI_ISL_985107 | 2021-01-07 | B.1.1.318 |
| hCoV-19/Nigeria/BCVL-18517/2021 | EPI_ISL_985117 | 2021-01-07 | B.1.525 |
| hCoV-19/Nigeria/BCVL-18556/2021 | EPI_ISL_985118 | 2021-01-07 | B.1.525 |
| hCoV-19/Nigeria/BCVL-18569/2021 | EPI_ISL_985119 | 2021-01-07 | B.1.525 |
| hCoV-19/Nigeria/BCVL-18317/2021 | EPI_ISL_985120 | 2021-01-07 | B.1.525 |
| hCoV-19/Nigeria/BCVL-18600/2021 | EPI_ISL_985121 | 2021-01-07 | B.1.525 |
| hCoV-19/Nigeria/BCVL-18623/2021 | EPI_ISL_985122 | 2021-01-07 | B.1.525 |
| hCoV-19/Nigeria/BCVL-18561/2021 | EPI_ISL_985127 | 2021-01-07 | B.1 |
| hCoV-19/Nigeria/BCVL-18758/2021 | EPI_ISL_985059 | 2021-01-08 | B.1.1.7 |
| hCoV-19/Nigeria/BCVL-18709/2021 | EPI_ISL_985082 | 2021-01-08 | B.1.1.7 |
| hCoV-19/Nigeria/BCVL-18716/2021 | EPI_ISL_985083 | 2021-01-08 | B.1.1.7 |
| hCoV-19/Nigeria/BCVL-18748/2021 | EPI_ISL_985084 | 2021-01-08 | B.1.1.7 |
| hCoV-19/Nigeria/BCVL-18387/2021 | EPI_ISL_985085 | 2021-01-08 | B.1.1.7 |
| hCoV-19/Nigeria/BCVL-18420/2021 | EPI_ISL_985086 | 2021-01-08 | B.1.1.7 |
| hCoV-19/Nigeria/BCVL-18423/2021 | EPI_ISL_985087 | 2021-01-08 | B.1.1.7 |
| hCoV-19/Nigeria/BCVL-18430/2021 | EPI_ISL_985088 | 2021-01-08 | B.1.1.7 |
| hCoV-19/Nigeria/BCVL-18458/2021 | EPI_ISL_985089 | 2021-01-08 | B.1.1.7 |
| hCoV-19/Nigeria/BCVL-18475/2021 | EPI_ISL_985090 | 2021-01-08 | B.1.1.7 |
| hCoV-19/Nigeria/BCVL-18757/2021 | EPI_ISL_985105 | 2021-01-08 | B.1.1.318 |
| hCoV-19/Nigeria/BCVL-18755/2021 | EPI_ISL_985114 | 2021-01-08 | B.1.525 |
| hCoV-19/Nigeria/BCVL-18762/2021 | EPI_ISL_985115 | 2021-01-08 | B.1.525 |
| hCoV-19/Nigeria/BCVL-18937/2021 | EPI_ISL_985116 | 2021-01-08 | B.1.525 |
| hCoV-19/Nigeria/BCVL-18760/2021 | EPI_ISL_985125 | 2021-01-08 | B.1.1.10 |
| hCoV-19/Nigeria/BCVL-18710/2021 | EPI_ISL_985129 | 2021-01-08 | B.1.1.10 |
| hCoV-19/Nigeria/BCVL-18756/2021 | EPI_ISL_985237 | 2021-01-08 | A |
| hCoV-19/Nigeria/BCVL-18684/2021 | EPI_ISL_985079 | 2021-01-09 | B.1.1.7 |
| hCoV-19/Nigeria/BCVL-18835/2021 | EPI_ISL_985080 | 2021-01-09 | B.1.1.7 |
| hCoV-19/Nigeria/BCVL-18483/2021 | EPI_ISL_985081 | 2021-01-09 | B.1.1.7 |
| hCoV-19/Nigeria/BCVL-18688/2021 | EPI_ISL_985073 | 2021-01-10 | B.1.1.7 |
| hCoV-19/Nigeria/BCVL-18798/2021 | EPI_ISL_985074 | 2021-01-10 | B.1.1.7 |
| hCoV-19/Nigeria/BCVL-18800/2021 | EPI_ISL_985075 | 2021-01-10 | B.1.1.7 |
| hCoV-19/Nigeria/BCVL-18820/2021 | EPI_ISL_985076 | 2021-01-10 | B.1.1.7 |
| hCoV-19/Nigeria/BCVL-18821/2021 | EPI_ISL_985077 | 2021-01-10 | B.1.1.7 |
| hCoV-19/Nigeria/BCVL-18949/2021 | EPI_ISL_985078 | 2021-01-10 | B.1.1.7 |
| hCoV-19/Nigeria/BCVL-18691/2021 | EPI_ISL_985111 | 2021-01-10 | B.1.525 |
| hCoV-19/Nigeria/BCVL-18828/2021 | EPI_ISL_985112 | 2021-01-10 | B.1.525 |
| hCoV-19/Nigeria/BCVL-18839/2021 | EPI_ISL_985113 | 2021-01-10 | B.1.525 |
| hCoV-19/Nigeria/BCVL-19040/2021 | EPI_ISL_985066 | 2021-01-11 | B.1.1.7 |
| hCoV-19/Nigeria/BCVL-19041/2021 | EPI_ISL_985067 | 2021-01-11 | B.1.1.7 |
| hCoV-19/Nigeria/BCVL-18975/2021 | EPI_ISL_985068 | 2021-01-11 | B.1.1.7 |
| hCoV-19/Nigeria/BCVL-18971/2021 | EPI_ISL_985069 | 2021-01-11 | B.1.1.7 |
| hCoV-19/Nigeria/BCVL-18976/2021 | EPI_ISL_985070 | 2021-01-11 | B.1.1.7 |
| hCoV-19/Nigeria/BCVL-18985/2021 | EPI_ISL_985071 | 2021-01-11 | B.1.1.7 |
| hCoV-19/Nigeria/BCVL-19053/2021 | EPI_ISL_985072 | 2021-01-11 | B.1.1.7 |
| hCoV-19/Nigeria/BCVL-19050/2021 | EPI_ISL_985128 | 2021-01-11 | B.1.1.10 |
| hCoV-19/Nigeria/BCVL-19097/2021 | EPI_ISL_985063 | 2021-01-12 | B.1.1.7 |
| hCoV-19/Nigeria/BCVL-19098/2021 | EPI_ISL_985064 | 2021-01-12 | B.1.1.7 |
| hCoV-19/Nigeria/BCVL-19205/2021 | EPI_ISL_985065 | 2021-01-12 | B.1.1.7 |
| hCoV-19/Nigeria/BCVL-20332/2021 | EPI_ISL_985062 | 2021-01-13 | B.1.1.7 |
| hCoV-19/Nigeria/BCVL-19703/2021 | EPI_ISL_985060 | 2021-01-14 | B.1.1.7 |
| hCoV-19/Nigeria/BCVL-19253/2021 | EPI_ISL_985061 | 2021-01-14 | B.1.1.7 |
| hCoV-19/Nigeria/BCVL-19702/2021 | EPI_ISL_985108 | 2021-01-14 | B.1.525 |
| hCoV-19/Nigeria/BCVL-19706/2021 | EPI_ISL_985109 | 2021-01-14 | B.1.525 |
| hCoV-19/Nigeria/BCVL-19707/2021 | EPI_ISL_985110 | 2021-01-14 | B.1.525 |
| hCoV-19/Nigeria/BCVL-19704/2021 | EPI_ISL_985126 | 2021-01-14 | B.1 |
