## Supplementary figures and images for "Coincident rapid expansion of two SARS-CoV-2 lineages with enhanced infectivity in Nigeria"

### Supplementary Table 2

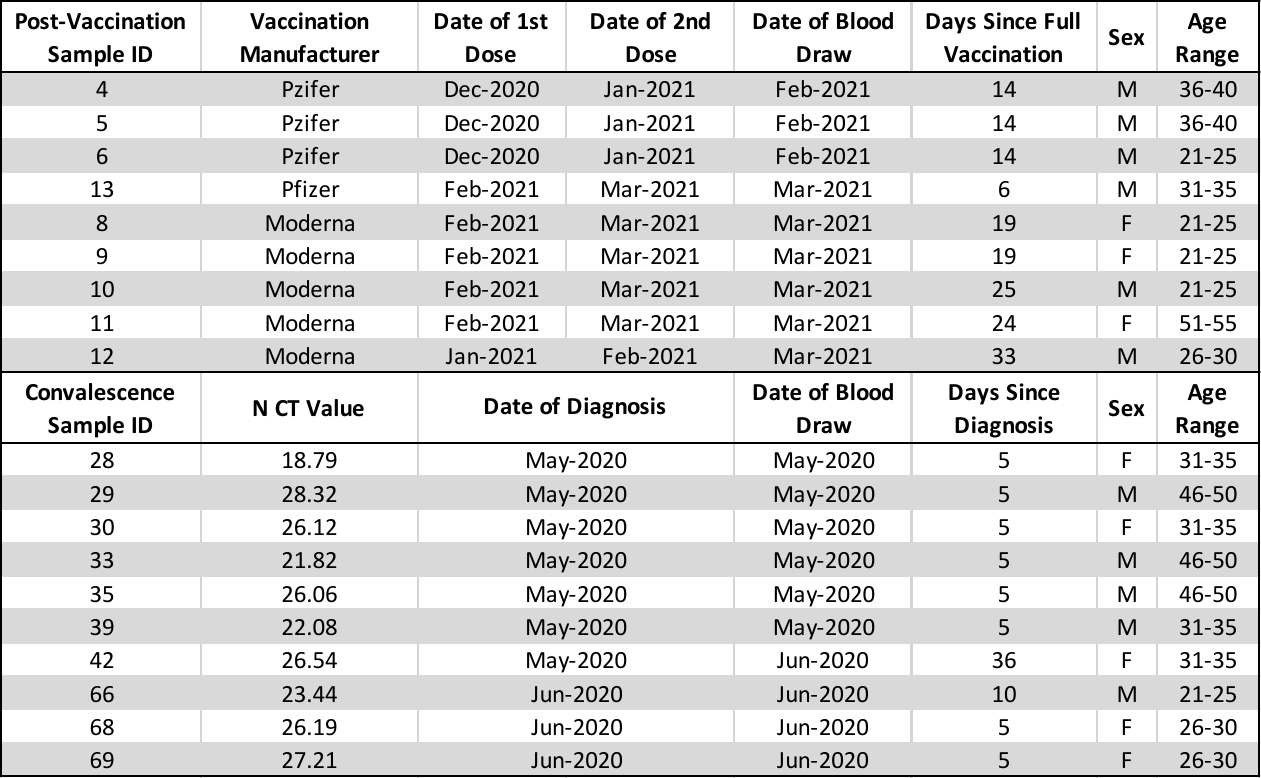
